# Comparative effectiveness of preventive strategies against medically-attended respiratory syncytial virus in U.S. infants during the first six months of life, 2023-2025

**DOI:** 10.64898/2026.08.25.26361361

**Authors:** Sara S Kim, Seth Zissette, Connor Van Meter, Machi Shiiba, Marina Bruck, Ashley Tippett, Satoshi Kamidani, David Benkeser, Elizabeth T Rogawski McQuade

## Abstract

**Key points:** *Question:* Among infants eligible to receive protection from maternal vaccination or infant long-acting monoclonal antibodies, what is the effectiveness RSV preventive strategies when accounting for real-world delays in administration?

*Findings:* In this target trial emulation study of 120,586 mother-infant pairs from U.S. claims data, long-acting monoclonal antibody effectiveness was sensitive to implementation delays, rendering preventive strategies that protected infants at or close to birth more effective at preventing respiratory disease than a monoclonal antibody strategy that reflected real-world implementation.

*Meaning:* Strategies, such as maternal vaccination or monoclonal antibodies provided within the first week of life, that protect infants at birth or close to birth are especially valuable in settings where delayed long-acting monoclonal antibody receipt is likely.

**Importance:** Maternal vaccination and long-acting monoclonal antibodies are now available in the U.S. to prevent RSV. Long-acting monoclonal antibody administration in the U.S. commonly occurs after hospital discharge in outpatient settings, leaving some infants unprotected early in life when severe RSV risk is highest. Comparative effectiveness between the two interventions and whether delays affect effectiveness estimates have not been quantified.

**Objective:** To evaluate the effectiveness of infant long-acting monoclonal antibody strategies and a maternal vaccination strategy, each compared to no intervention, and the comparative effectiveness of intervention strategies when accounting for real-world delays in monoclonal antibody receipt.

**Design:** Cohort study using target trial emulation to compare four strategies for prevention of RSV-related outcomes.

**Setting:** The U.S. between 2023 and 2025 using a nationwide database of employer-sponsored commercial insurance claims.

**Participants:** 120,586 commercially insured mother-infants, whose infants were born in the U.S. during the 2023-2024 or 2024-2025 RSV season. Infants who could not be paired with their mother’s record, did not enroll in commercial insurance within 75 days from birth, received palivizumab, and had an implausible birth date were excluded.

**Interventions:** Comparison of four RSV prevention strategies: (i) maternal RSVpreF; (ii) long-acting monoclonal antibody given within the first week of life (mAb as intended); (iii) long-acting monoclonal antibody given within a six-month grace period from birth (mAb within grace period); and (iv) a control.

**Main outcomes and measures:** Effectiveness against first RSV-associated hospitalization and medically-attended RSV illness was summarized using adjusted hazard ratios (aHR) and estimated using an inverse propensity weighting approach, with weights accounting for maternal age, maternal comorbidities affecting pregnancy, obstetric and newborn complications, season, region, and birth timing relative to October 1. A weighted Kaplan Meier estimator was used to estimate strategy-specific cumulative incidence of RSV outcomes over time.

**Results:** In the first five weeks of life, the mAb within grace period strategy doubled the hazard of RSV hospitalization (aHR: 2.0 [95% CI: 1.0-4.9]) and increased the hazard of medically-attended RSV (aHR: 1.6 [95% CI: 1.0-2.7]) compared to the maternal RSVpreF strategy. The hazard for RSV hospitalization was similar for the mAb as intended strategy compared to the maternal RSVpreF strategy (aHR = 0.9 [95% CI: 0.3-1.9]).

**Conclusions and relevance:** RSVpreF and monoclonal antibodies were similarly effective when monoclonal antibodies were administered close to birth, but when accounting for real-world delays in monoclonal antibody receipt, the maternal RSVpreF strategy was more effective than the mAb within grace period strategy.

## Introduction

Respiratory syncytial virus (RSV) is the leading cause of infant hospitalization in the United States (U.S.).^1^ Among U.S. children aged less than five years, an estimated 2.1 million children have an RSV-related medically-attended illness annually.^2,3^

In 2023, the U.S. licensed a maternal vaccine and long-acting monoclonal antibody (mAb) as RSV preventive interventions for infants. The bivalent prefusion F (RSVpreF) vaccine is recommended during pregnancy at 32-36 weeks for those expecting to deliver during the RSV season.^4^ mAbs are recommended for infants under eight months entering their first RSV season and whose mothers did not receive RSVpreF.^5^ Infants born during the RSV season are recommended to receive mAbs during the first week of life.^6^ Infants expected to be born during the RSV season may be protected through either maternal RSVpreF or mAbs.

Two French studies have directly compared these interventions, observing that infant mAb receipt was associated with a lower risk of RSV-related lower respiratory tract hospitalization compared to maternal vaccination.^7,8^ Importantly, these analyses were limited to infants receiving mAbs during birth hospitalization and assessed protection from receipt, consistent with prior mAb studies.^9–18^ However, mAb administration in the U.S. commonly occurs after hospital discharge in outpatient settings,^14^ leaving some infants unprotected early in life when severe RSV risk is highest.^19^ Whether delays affect mAb effectiveness has not been quantified.

Using a large database of privately insured individuals born during the 2023-2024 and 2024-2025 RSV seasons, we estimate 1) the effectiveness of maternal RSVpreF and mAb strategies received before six months of age, each versus no intervention, and 2) the effectiveness of mAb strategies versus a maternal RSVpreF strategy, against RSV, bronchiolitis, and all-cause acute respiratory illness (ARI) associated outcomes through six months of age among U.S. infants. We account for real-world mAb administration delays^20^ to better inform RSV prevention decisions in U.S. healthcare systems. To isolate the impact of mAb administration timing on effectiveness, we compare RSV risk under a scenario that reflects observed delays in administration with one that enforces mAb receipt during the first week of life.

## Methods

### Data source

Using a previously published linkage algorithm,^21^ we constructed a mother-infant birth cohort in MarketScan, a U.S.-based administrative claims database of commercially insured individuals and their dependents (Appendix S1).^22^ Data included diagnosis codes, procedures, and dates related to inpatient and outpatient medical encounters, insurance enrollment status, and basic demographics.

### Study design

We conducted a cohort study using target trial emulation to estimate the effectiveness of preventive strategies during the infant’s first six months of life (Table S1). We followed infant outcomes at weekly intervals from birth until the first occurrence of an outcome, insurance lapse during follow-up, completion of six months of follow-up, or deviation from assigned intervention strategy.

### Eligibility

We included infants born during the 2023-2024 and 2024-2025 RSV seasons (October 1-March 31). We excluded infants unable to be paired with their mother’s record, not enrolled in commercial insurance within 75 days from birth, who received palivizumab, and whose birth date could not be reliably inferred.

### Intervention strategies

We considered estimation of risk of RSV outcomes under four hypothetical RSV prevention strategies: (i) maternal RSVpreF; (ii) mAb as intended; (iii) mAb within grace period; and (iv) a control.

The maternal RSVpreF strategy reflects the current recommendations for RSVpreF vaccination: mothers are recommended to receive RSVpreF during the third trimester of pregnancy; however, infants born within 14 days of vaccination are recommended to also receive mAbs. This strategy estimates the effectiveness of a policy primarily based on maternal vaccination but allowing for mAbs for infants born shortly after vaccination (Table S2).

The mAb as intended strategy evaluates the impact of mAb receipt within the first week of life, as intended by policy. This timing reflects a gold standard of implementation, though may not be universally feasible in real-world settings. Infants who received mAbs after the first week of life contribute follow up time to this strategy from birth until an RSV-associated outcome, right-censoring event, or end of the first week of life, whichever occurs first (Table S2).

Under the mAb within grace period strategy, infants born in an RSV season can receive mAbs anytime within six months from birth. The timing of mAb administration over this period matches real-world patterns of mAb uptake and healthcare delivery and aligns with national guidelines within the follow-up period of interest. Infants who received mAbs outside of the grace period contribute follow up time until an RSV-associated outcome or right-censoring event, whichever occurs first (Table S2). Infants who received mAbs as intended also received mAbs in alignment with this strategy; thus, these strategies are not mutually exclusive. Comparison of outcomes under this strategy to the mAb as intended strategy quantifies the impact of real-world delays in mAb administration.

The control condition required receiving neither maternal RSVpreF nor mAbs before six months of age.

We used Current Procedural Terminology (CPT) codes in both inpatient and outpatient settings for RSV preventive products (Appendix S2) to classify the weekly periods after infant birth that aligned with each of the prevention strategies.

### Outcome

Primary outcomes were first RSV-associated hospitalization defined by an inpatient encounter with an RSV International Classification of Diseases, 10^th^ Revision (ICD-10) code (J21.0, J12.1, J20.5, J21.9) and first medically-attended RSV illness defined by an inpatient or outpatient encounter with an RSV ICD-10 code. Outpatient encounters included office, emergency department, and urgent care visits.

Secondary outcomes included first bronchiolitis hospitalization, medically-attended bronchiolitis, ARI hospitalization, and medically-attended ARI (Appendix S3), as well as a stricter RSV definition requiring both an RSV ICD-10 code and an RSV testing CPT code (Appendix S4). RSV outcomes were not necessarily a subset of the broader outcomes, nor bronchiolitis within ARI, given the inclusion of first episode only.

### Analysis

We characterized the effectiveness of intervention strategies using a pooled logistic marginal structural model (MSM) that approximated discrete-time adjusted hazard ratios (aHR). We used inverse propensity weighting to estimate the parameters of the MSM. The weights required propensity models describing (i) the probability of RSVpreF during pregnancy given maternal characteristics; (ii) the time-varying probability of mAb initiation after birth given maternal characteristics, maternal receipt of RSVpreF and timing of receipt, and newborn characteristics; (iii) the time-varying probability of right-censoring after birth due to insurance disenrollment. Maternal characteristics considered included maternal age, maternal comorbidities affecting pregnancy (Appendix S5), number of prenatal healthcare encounters during pregnancy, and obstetric complications.^23^ Newborn characteristics included newborn complications including preterm birth, congenital circulatory malformations and pulmonary abnormalities (Appendix S6), birth season, birth region, and a cross-product between age and birth month relative to October 1.^9,24^ These probabilities were separately estimated using pooled logistic regression.

The estimated propensities were used to construct weekly weights corresponding to each prevention strategy. We cloned mother-infant pairs across intervention strategies that had non-zero weights and estimated the parameters of a pooled logistic MSM by regressing the outcome of interest on the intervention strategy and a third-degree polynomial for week since birth in the cloned dataset.

We calculated prevention efficacy relative to control as 1 exp 100%, where represents the coefficient for the active prevention intervention in the MSM. For comparing active prevention strategies to each other, we report aHRs rather than prevention efficacy.

We assessed effect modification according to RSV season, preterm birth status, born early or late in the season, region, and follow-up period (≤5 weeks and >5 weeks) by using separate MSMs that each included a cross-product between the relevant effect modifier and intervention strategy.

In addition to aHRs, we used the same inverse propensity weights to estimate strategy-specific cumulative incidence of RSV outcomes over time using a weighted Kaplan Meier estimator. Cumulative risk differences (RD) for each contrast were calculated weekly from birth to six months of age. These estimates were used to report the estimated number needed to immunize (NNI) to reduce incidence by one case in comparison to a reference strategy.

We reported 95% confidence intervals (CIs) using a percentile-based nonparametric bootstrap based on 1000 bootstrap samples. When the associated RD 95% CI crossed zero, we presented discontinuous intervals for NNI.^25^ Analyses were conducted using R version 4.4.2.

The protocol and study procedures were reviewed and approved by the Institutional Review Board at Emory University (Protocol #:2025P013207).

## Results

We included 120,586 infants (81%) among 148,051 infants born during the 2023-2024 or 2024-2025 RSV seasons (n=23,081 not paired with mother’s record, n=4301 not enrolled within 75 days from birth, n=82 received palivizumab, n=1 had implausible birth date).

Over both seasons, 19,703 (16%) mothers received RSVpreF during pregnancy (Table 1) with lower uptake in 2023-2024 (8%) than 2024-2025 (25%). Infants born December-February were more likely than those born earlier in the season to have vaccinated mothers, and uptake differed by region (Table 1). Among 882 infants born within 14 days of maternal RSVpreF, 229 (26%) subsequently received mAbs before six months of age (Table S3). Among 18,821 infants born ≥14 days after maternal RSVpreF, 551 (3%) also received mAbs and were censored from both strategies at mAb receipt.

**Table 1.** Descriptive characteristics by observed intervention strategy^a^.

|  | Maternal<br>RSVpreF | mAb as<br>intended | mAb within<br>grace period | Control |
| --- | --- | --- | --- | --- |
|  | N = 19,703<sup>b</sup> | N = 7227<sup>b</sup> | N = 18,388<sup>b</sup> | N = 83,046<sup>b</sup> |
| **RSV Hospitalization** | 41 (0.2%) | 11 (0.2%) | 54 (0.3%) | 661 (0.8%) |
| **Medically-Attended RSV** | 229 (1.2%) | 97 (1.3%) | 358 (1.9%) | 2230 (2.7%) |
| **Birth Season** |  |  |  |  |
| 2023-2024 | 5062 (26%) | 2838 (39%) | 8907 (48%) | 49,260 (59%) |
| 2024-2025 | 14,641 (74%) | 4389 (61%) | 9481 (52%) | 33,786 (41%) |
| **Birth Month** |  |  |  |  |
| October | 2331 (12%) | 1221 (17%) | 5727 (31%) | 14,504 (17%) |
| November | 3045 (15%) | 1382 (19%) | 3815 (21%) | 13,148 (16%) |
| December | 3629 (18%) | 1207 (17%) | 2731 (15%) | 11,925 (14%) |
| January | 4429 (22%) | 1234 (17%) | 2643 (14%) | 13,732 (17%) |
| February | 3943 (20%) | 1100 (15%) | 1914 (10%) | 13,162 (16%) |
| March | 2326 (12%) | 1083 (15%) | 1558 (8.5%) | 16,575 (20%) |
| **Maternal Age** | 32.81 (4.33) | 32.25 (4.42) | 32.23 (4.47) | 32.18 (4.55) |
| **Number of Prenatal Encounters** | 4.48 (3.24) | 4.16 (3.17) | 4.14 (3.17) | 4.04 (3.17) |
| **Birth Region** |  |  |  |  |
| Midwest | 5878 (30%) | 1770 (25%) | 4264 (24%) | 15,071 (18%) |
| Northeast | 3740 (19%) | 1140 (16%) | 2786 (15%) | 14,311 (17%) |
| South | 5207 (27%) | 2827 (40%) | 7591 (42%) | 37,721 (46%) |
| West | 4515 (23%) | 1336 (19%) | 3450 (19%) | 15,027 (18%) |
| Unknown | 363 | 154 | 297 | 916 |
| **Preterm Birth** | 1617 (8.2%) | 653 (9.0%) | 2174 (12%) | 9533 (11%) |
| **Any Newborn Complication** | 5104 (26%) | 1806 (25%) | 5460 (30%) | 24,874 (30%) |
| **Maternal Comorbidity Affecting Pregnancy** | 13,086 (66%) | 4622 (64%) | 11,743 (64%) | 52,047 (63%) |
| **Maternal Obstetric Complication** | 9714 (49%) | 3370 (47%) | 9082 (49%) | 40,242 (48%) |

<sup>a</sup>Preventive intervention strategies are not mutually exclusive. All infants in the mAb as intended strategy are a subset of infants in the mAb within grace period strategy. Additionally, infants who received mAbs may have had mothers who received maternal RSVpreF. See supplementary tables S2-S4.
<sup>b</sup>n (%); Mean (Standard Deviation).
Abbreviations: mAb: long-acting monoclonal antibody; RSVpreF: Bivalent prefusion F vaccine

Within six months from birth, 18,388 (15%) infants received mAbs (Table 1). Among those infants, 39% received them in the first week of life. Infants born earlier in the season (October-December) were more likely than infants born later to receive mAbs. Among infants receiving mAbs after the first week of life (Table S4), median age at receipt was 22 days (interquartile range (IQR): 14-40). Infants received mAbs sooner in 2024-2025 (median: 8 days (IQR: 4-19)) compared to 2023-2024 (median: 23 days (IQR: 6-69) (Figure S1).

Infants who received mAbs as intended and whose mothers received RSVpreF were less likely to be born prematurely or have a newborn complication than infants who received mAbs within the grace period but after the first week of life (Table 1).

Infants born within 14 days of maternal RSVpreF who also received mAbs were more likely to have newborn complications and mothers with comorbidities or obstetric complications than infants in all other intervention strategies (Table S3).

### Maternal RSVpreF vaccination effectiveness

In the first six months of life, the maternal RSVpreF strategy reduced the hazard of RSV hospitalization by 83.1% (95% CI: 76.1%-89.3%) and medically-attended RSV by 65.8% (95% CI: 59.0%-71.6%) versus no intervention (Table 2). Estimates for both RSV outcomes were higher in 2023-2024 than 2024-2025 and in the western region versus elsewhere. Among term infants, effectiveness was higher against RSV hospitalization. At six months, this strategy reduced the absolute risk of RSV hospitalization by 68.7 cases (95% CI: 60.2-78.3) per 10,000 infants, for an NNI of 145 (95% CI: 127-166) and reduced medically-attended RSV by 214.4 cases (95% CI: 187.8-238.7) per 10,000 infants, for an NNI of 46 (95% CI: 41-53) (Table 3). While only 46% of medically-attended RSV encounters and 29% of RSV hospitalizations had an RSV test ordered, effectiveness was similar when restricted to tested outcomes (Table S5).

**Table 2.** Summary effectiveness ([1-aHR]*100%) of intervention strategy compared to control.

|  | **Maternal RSVpreF vs control** | **mAb as intended vs control** | **mAb within grace period vs control** |
| --- | --- | --- | --- |
| **RSV hospitalization** | 83.1% (76.1% - 89.3%) | 84.6% (73.1% - 93.8%) | 79.0% (70.8% - 86.3%) |
| **Season** |  |  |  |
| 2023-2024 | 97.2% (90.8% - 100.0%) | 91.9% (79.7% - 100.0%) | 75.3% (64.0% - 85.6%) |
| 2024-2025 | 66.3% (52.1% - 79.3%) | 76.5% (56.1% - 92.8%) | 82.2% (69.8% - 92.3%) |
| **Preterm** |  |  |  |
| No | 84.1% (76.4% - 90.8%) | 86.4% (75.2% - 96.0%) | 79.5% (70.7% - 87.4%) |
| Yes | 74.9% (54.8% - 94.8%) | 68.3% (17.4% - 100.0%) | 75.1% (49.4% - 93.0%) |
| **Birth relative to season** |  |  |  |
| Early (Oct – Dec) | 78.2% (68.0% - 87.1%) | 82.0% (67.9% - 93.0%) | 77.2% (67.6% - 85.5%) |
| Late (Jan – Mar) | 87.9% (77.4% - 91.8%) | 90.1% (65.7% - 100.0%) | 81.9% (60.7% - 95.8%) |
| **Region** |  |  |  |
| Midwest | 87.6% (78.4% - 95.5%) | 87.9% (68.9% - 100.0%) | 87.9% (77.0% - 96.2%) |
| Northeast | 67.8% (34.2% - 90.6%) | 74.6% (34.6% - 100.0%) | 82.6% (58.5% - 98.5%) |
| South | 80.7% (66.8% - 93.5%) | 76.1% (53.0% - 94.6%) | 65.1% (47.5% - 80.7%) |
| West | 90.8% (82.4% - 97.1%) | 100.0% (100.0% - 100.0%) | 88.0% (76.4% - 96.2%) |
| **Medically-attended RSV** | 65.8% (59.0% - 71.6%) | 66.0% (57.7% - 74.2%) | 58.0% (51.2% - 63.7%) |
| **Season** |  |  |  |
| 2023-2024 | 86.0% (79.3% - 92.1%) | 84.4% (75.8% - 91.6%) | 65.6% (57.7% - 72.6%) |
| 2024-2025 | 46.0% (34.0% - 56.3%) | 51.3% (36.6% - 63.3%) | 51.8% (40.1% - 60.9%) |
| **Preterm** |  |  |  |
| No | 65.2% (57.4% - 71.7%) | 66.8% (58.5% - 74.7%) | 58.7% (51.7% - 64.9%) |
| Yes | 72.1% (55.7% - 85.2%) | 57.6% (23.1% - 85.6%) | 52.1% (27.8% - 71.0%) |
| **Birth relative to season** |  |  |  |
| Early (Oct – Dec) | 64.0% (57.1% - 70.4%) | 69.0% (60.8% - 76.8%) | 60.4% (53.6% - 66.4%) |
| Late (Jan – Mar) | 46.1% (20.0% - 67.1%) | 43.2% (14.8% - 67.1%) | 37.3% (12.5% - 57.4%) |
| **Region** |  |  |  |
| Midwest | 63.7% (51.5% - 73.6%) | 80.4% (68.6% - 91.8%) | 73.8% (63.8% - 82.5%) |
| Northeast | 57.8% (40.3% - 72.8%) | 61.2% (35.4% - 84.8%) | 58.2% (38.7% - 75.6%) |
| South | 62.3% (49.7% - 72.9%) | 56.0% (42.5% - 69.1%) | 47.2% (36.8% - 57.0%) |
| West | 81.4% (70.8% - 89.4%) | 74.4% (55.7% - 88.7%) | 62.9% (47.1% - 76.6%) |

Abbreviations: aHR: adjusted hazard ratio; mAb: long-acting monoclonal antibody; RSVpreF:
Bivalent prefusion F vaccine

**Table 3:** Risk difference per 10,000 infants and number needed to immunize with 95% confidence intervals of outcomes by intervention strategy over six months since birth.

|  | Maternal RSVpreF vs control | mAb as intended vs control | mAb within grace period vs control | mAb within grace period vs mAb as intended | mAb as intended vs Maternal RSVpreF | mAb within grace period vs Maternal RSVpreF |
| --- | --- | --- | --- | --- | --- | --- |
| **PRIMARY OUTCOMES** |  |  |  |  |  |  |
| **RSV hospitalization** |  |  |  |  |  |  |
| Risk difference | -68.7 (-78.3 - -60.2) | -69.5 (-80.5 - -58.1) | -65.3 (-74.4 - -56.2) | 4.2 (-1.5 - 9.5) | -0.8 (-10.6 - 9.6) | 3.5 (-5.0 - 12.3) |
| NNI | 145 (127 - 166) | 143 (124 - 172) | 153 (134 - 177) | 2353 (NNTB: 6528 - ∞, NNTH: 1054 - ∞) | 12,969 (NNTB: 943 - ∞, NNTH: 1042 - ∞)<sup>a</sup> | 2874 (815 - 2012) |
| **Medically attended RSV** |  |  |  |  |  |  |
| Risk difference | -214.3 (-238.7 - -187.8) | -215.0 (-244.5 - -185.5) | -189.7 (-213.7 - -164.6) | 25.3 (9.9 - 41.5) | -0.7 (-35.2 - 34.3) | 24.5 (-6.2 - 55.5) |
| NNI | 46 (41 - 53) | 46 (40 - 53) | 52 (46 - 60) | 395 (241 - 1006) | 13,875 (NNTB: 284 - ∞, NNTH: 291 - ∞)<sup>a</sup> | 407 (NNTB: 1608 - ∞, NNTH: 180 - ∞)<sup>a</sup> |
| **SECONDARY OUTCOMES** |  |  |  |  |  |  |
| **Bronchiolitis hospitalization** |  |  |  |  |  |  |
| Risk difference | -187.5 (-358.9 - -24.1) | -68.1 (-332.0 - 199.0) | -92.6 (-294.3 - 109.6) | -24.5 (155.5 - 86.3) | 119.4 (-185.1 - 0.0) | 94.9 (-149.7 - 334.5) |
| NNI | 53 (28 - 414) | 147 (NNTB: 30 - ∞, NNTH: 50 - ∞)<sup>a</sup> | 108 (NNTB: 33 - ∞, NNTH: 91 - ∞)<sup>a</sup> | 408 (NNTB: 63 - ∞, NNTH: 116 - ∞)<sup>a</sup> | 83 (NNTB: 54 - ∞, NNTH: 23 - ∞)<sup>a</sup> | 105 (NNTB: 66 - ∞, NNTH: 29 - ∞)<sup>a</sup> |
| **Medically-attended bronchiolitis** |  |  |  |  |  |  |
| Risk difference | -1031.8 (-1371.0 - -675.1) | -720.1 (-1142.7 - -290.9) | -571.3 (-914.9 - -193.6) | 149.2 (-98.8 - 405.6) | 311.3 (-236.1 - 819.5) | 460.5 (-8.05 - 893.1) |
| NNI | 9 (7 - 14) | 13 (8 - 34) | 17 (10 - 51) | 67 (NNTB: 101 - ∞, NNTH: 24 - ∞)<sup>a</sup> | 32 (NNTB: 42 - ∞, NNTH: 12 - ∞)<sup>a</sup> | 21 (NNTB: 1241 - ∞, NNTH: 11 - ∞)<sup>a</sup> |
| **ARI hospitalization** |  |  |  |  |  |  |
| Risk difference | -291.6 (-498.3 - -60.0) | -238.5 (-552.5 - 62.2) | -219.1 (-456.7 - 20.1) | 19.5 (-134.7 - 173.3) | 53.1 (-303.2 - 390.6) | 72.5 (-213.9 - 364.6) |
| NNI | 34 (20 - 166) | 41 (NNTB: 19 - ∞, NNTH: 160 - ∞)<sup>a</sup> | 45 (NNTB: 22 - ∞, NNTH: 498 - ∞)<sup>a</sup> | 513 (NNTB: 74 - ∞, NNTH: 57 - ∞)<sup>a</sup> | 188 (NNTB: 32 - ∞, NNTH: 25 - ∞)<sup>a</sup> | 137 (NNTB: 46 - ∞, NNTH: 27 - ∞)<sup>a</sup> |
| **Medically-attended ARI** |  |  |  |  |  |  |
| Risk difference | -124.1 (-188.0 - -61.5) | 19.0 (-33.7 - 65.2) | 1.2 (-43.2 - 39.0) | -17.8 (-41.8 - 12.2) | 143.1 (59.8 - 218.4) | 125.3 (54.4 - 196.2) |
| NNI | 80 (53 - 162) | 527 (NNTB: 297 - ∞, NNTH: 153 - ∞)<sup>a</sup> | 8487 (NNTB: 231 - ∞, NNTH: 256 - ∞)<sup>a</sup> | 562 (NNTB: 239 - ∞, NNTH: 821 - ∞)<sup>a</sup> | 69 (45 - 167) | 8487 (50 - 183) |

Abbreviations: mAb: long-acting monoclonal antibody; RSVpreF: Bivalent prefusion F vaccine; NNI: Number needed to immunize, NNTB: Number needed to benefit, NNTH: Number needed to harm
<sup>a</sup>Confidence intervals for number needed to immunize presented as discontinuous intervals when the 95% CI for the associated risk difference crosses the null according to Altman 1998.

The maternal RSVpreF strategy also provided protection against secondary outcomes (Table S6). At six months, this strategy reduced the absolute risk of bronchiolitis hospitalization by 187.5 cases (95% CI: 24.1-358.9) per 10,000 infants for an NNI of 53 (95% CI: 28-414) and medically-attended bronchiolitis by 1031.8 cases (95% CI: 675.1-1371.0) per 10,000 infants for an NNI of 9 (95% CI: 7-14) (Table 3). This strategy reduced the absolute risk of ARI hospitalization by 291.6 cases (95% CI: 60.0-498.3) per 10,000 infants and medically-attended ARI by 124.1 cases (95% CI: 61.5-199.0) per 10,000 infants in the first six months of life.

### Long-acting monoclonal antibody effectiveness

The mAb as intended strategy reduced the hazard of RSV hospitalization by 84.6% (95% CI: 73.1%-93.8%) and medically-attended RSV by 66.0% (95% CI: 57.7%-74.2%) versus no intervention over six months from birth (Table 2). Effectiveness for both RSV outcomes was higher in 2023-2024 than 2024-2025 and among term versus preterm infants but lower for northeastern and southern regions than elsewhere. At six months of follow-up, this strategy reduced the absolute risk of RSV hospitalization by 69.5 cases (95% CI: 58.1-80.5) per 10,000 infants (NNI: 143 [95% CI: 124-172]), medically-attended RSV by 215.0 cases (95% CI: 185.5-244.5) per 10,000 infants (NNI: 46 [95% CI: 40-53]), and medically-attneded bronchiolitis by 720.1 cases (95% CI: 290.9-1142.7) per 10,000 infant (NNI: 13 [95% CI: 8-34]) (Table 3).

The mAb within grace period strategy reduced the hazard of RSV hospitalization by 79.0% (95% CI: 70.8%-86.3%) and medically-attended RSV by 58.0% (95% CI: 51.2%-63.7%) versus no intervention in the first six months of life (Table 2).

Effectiveness against both RSV outcomes was lower in the south versus other regions and against medically-attended RSV was higher among infants born earlier in the season. This strategy reduced the six-month incidence of RSV hospitalization by 65.3 cases (95% CI: 56.2-74.4) per 10,000 infants, for an NNI of 153 (95% CI: 134-177) and medically-attended RSV by 189.7 cases (95% CI: 164.6-213.7) per 10,000 infants, for an NNI of 52 (95% CI: 46-60) (Table 3). This strategy reduced the absolute risk of medically-attended bronchiolitis by 571.3 cases (95% CI: 193.6-914.9) per 10,000 infants for an NNI of 17 (95% CI: 10-51) (Table 3). Effectiveness against RSV outcomes with a diagnostic test was similar to estimates using the primary outcome definitions (Table S5).

Compared to mAb as intended, the grace period strategy had a higher hazard for RSV hospitalization (aHR: 1.3 [95% CI: 0.9-2.5]), though not statistically significant, and medically-attended RSV (aHR: 1.2 [95% CI: 1.1-1.4]) (Table 4). This elevated hazard was sustained over follow-up (Figure S2) and particularly evident for RSV hospitalization in the first five weeks of life (aHR: 1.8 [95% CI: 1.1-5.7]) (Table S7). At six months, the grace period strategy increased the absolute risk of RSV hospitalization by 4.2 cases (95% CI: −1.5 −9.5) per 10,000 infants and medically-attended RSV by 25.3 cases (95% CI: 9.9-41.5) per 10,000 infants versus the as intended strategy (Table 3).

**Table 4.** Comparative effectiveness (adjusted hazard ratio) with 95% confidence interval of mAb within grace period versus mAb as intended strategy, mAb as intended versus maternal RSVpreF strategy, and mAb within grace period vs maternal RSVpreF strategy.

|  | **mAb within grace period vs mAb as intended** | **mAb as intended vs maternal RSVpreF** | **mAb within grace period vs maternal RSVpreF** |
| --- | --- | --- | --- |
| **RSV hospitalization** | 1.3 (0.9 – 2.5) | 0.9 (0.3 – 1.9) | 1.2 (0.7 – 2.2) |
| **Medically-attended RSV** | 1.2 (1.1 – 1.4) | 1.0 (0.7 -1.3) | 1.2 (1.0 -1.6) |

Abbreviations: mAb: long-acting monoclonal antibody; vs: versus; RSVpreF: Bivalent prefusion F vaccine

### Comparative effectiveness of mAbs versus maternal RSVpreF

The hazard for RSV hospitalization was similar for the mAb as intended strategy compared to the maternal RSVpreF strategy (aHR = 0.9 [95% CI: 0.3-1.9]) (Table 4). The weekly cumulative incidences for the two strategies were similar throughout follow-up (Figure 1).

**Figure 1.**
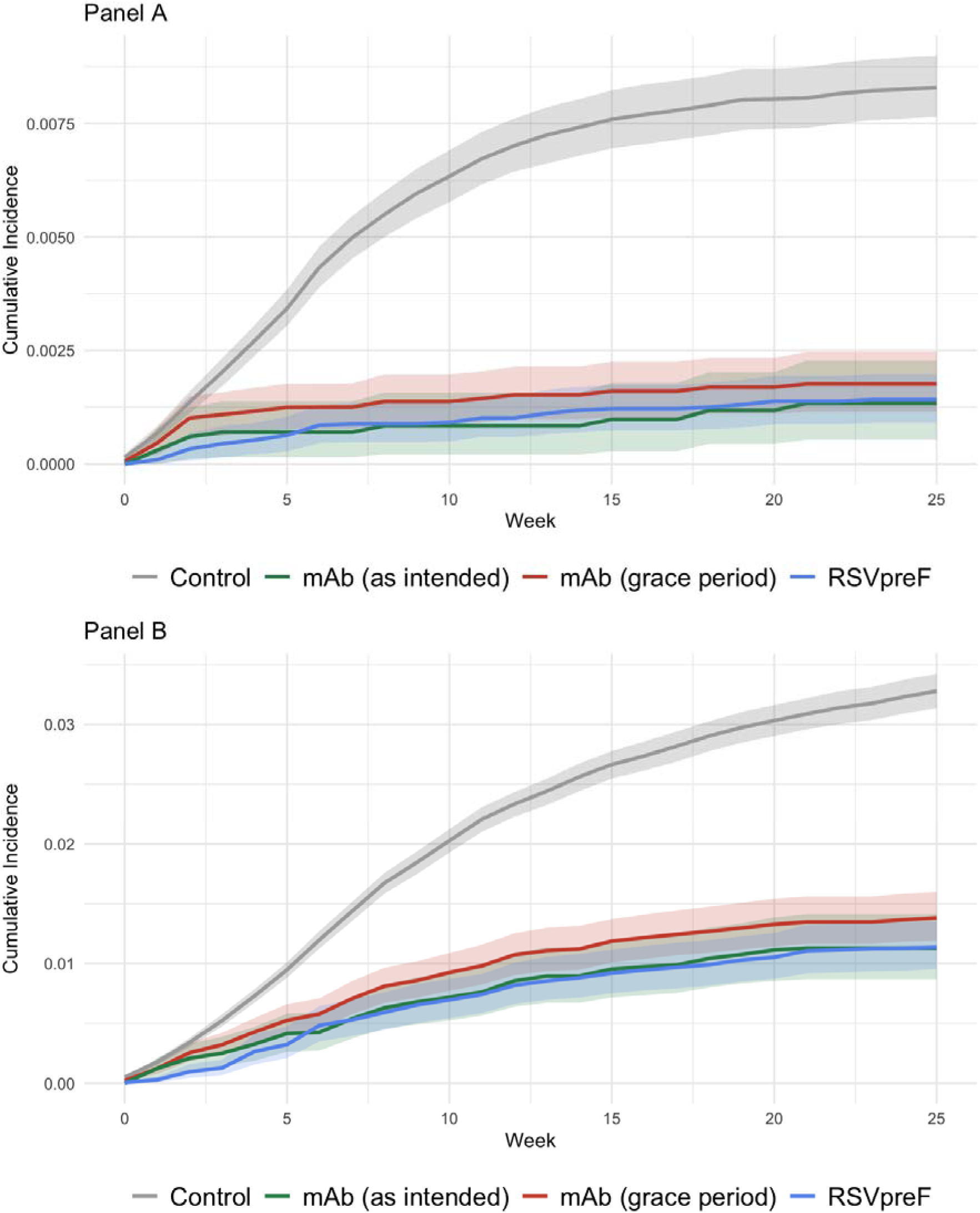
Cumulative incidence with 95% confidence intervals of RSV hospitalization and medically-attended RSV over time by intervention strategy. Caption: Panel A: Cumulative incidence of RSV hospitalization; Panel B: Cumulative incidence of medically-attended RSV Abbreviations: mAb: long-acting monoclonal antibody; RSVpreF: Bivalent prefusion F vaccine.

In the first five weeks of life, the mAb within grace period strategy doubled the hazard of RSV hospitalization compared to the maternal RSVpreF strategy (aHR: 2.0 [95% CI: 1.0-4.9]) and increased the hazard of medically-attended RSV (aHR: 1.6 [95% CI: 1.0-2.7]) (Table S7). This early divergence attenuated over the six-month follow-up for both RSV outcomes (Table 4). At six months, the mAb within grace period strategy increased the absolute risk of RSV hospitalization by 3.5 cases (95% CI: −5.0-12.3) per 10,000 infants and medically-attended RSV by 24.5 cases (95% CI: −6.2-55.5) per 10,000 infants (Table 3).

## Discussion

Maternal RSVpreF and mAb strategies reduced respiratory disease morbidity, but mAb effectiveness was sensitive to implementation delays. Our results suggest that mAb-based strategies that fail to achieve universal coverage in the first week of life will yield higher RSV hospitalization rates than strategies providing protection close to birth. The effectiveness gap likely reflects high early RSV risk prior to administration rather than any lasting difference in protection once mAbs are administered.

All intervention strategies provided large absolute risk reductions across RSV-specific and broader respiratory outcomes (Table 3). NNIs for medically-attended RSV (46-52) and RSV hospitalization (143-153) were comparable to those for medically-attended influenza and substantially lower than for influenza hospitalization, which requires an estimated 1000 vaccinations to reduce one hospitalization.^26^ NNIs for medically-attended bronchiolitis were particularly low: 9 (95% CI: 7-14) for the maternal RSVpreF strategy, 13 (95% CI: 8-34) for the mAb as intended strategy, and 17 (95% CI: 10-51) for the mAb within grace period strategy. These large all-cause bronchiolitis and ARI effects suggest frequent RSV undertesting and underdiagnosis such that RSV-specific effects underestimate the true value of these products. RSV causes an estimated 60-80% of bronchiolitis,^27^ but only 33% of medically-attended bronchiolitis cases were tested in this population. Absolute effects on non-specific respiratory outcomes may better capture the overall clinical burden of preventable RSV than estimates restricted to laboratory-confirmed outcomes, including those based on test-negative designs.

Effectiveness of mAbs and maternal RSVpreF was generally lower in 2024-2025 versus 2023-2024. Although RSVpreF recipients had similar measured characteristics across seasons (Table S8), uptake was low during the first season^28,29^ and unmeasured differences between earlier and later adopters may exist. Differences in maternal RSVpreF timing relative to RSV circulation also may have contributed. Although maternal vaccination uptake occurred earlier in 2024-2025 (Table S8), the season began later than in 2023-2024,^30^ and waning may have lowered effectiveness. Our 2024-2025 maternal RSVpreF effectiveness estimates were within range of other U.S.-based studies.^10,18,31^

mAb administration timing may also have influenced seasonal differences in effectiveness. First, effectiveness among mAb recipients dosed in the first week of life was lower in 2024-2025 (Table 2), possibly reflecting waning protection given the later season onset.^30,32^ Second, effectiveness for the mAb within grace period strategy against RSV hospitalization was higher in 2024-2025 (Table 2, Figure S3 Panel A-B), consistent with more timely mAb receipt that season compared to 2023-2024 (Figure S1).

This study is subject to limitations. Using claims data, we assumed that absence of a diagnosis or procedure indicated an event did not occur. Moreover, RSV outcomes were identified using ICD-10 codes, which have moderate sensitivity among children <5 years.^33^ Next, we could not exclude the possibility of residual confounding. Finally, because the study population consisted of commercially insured infants born during the RSV season, our findings may not be generalizable to wider populations or comparable to other mAb effectiveness studies.

Delayed mAb administration diminished real-world protection. When accounting for delays in mAb receipt, the maternal RSVpreF strategy was more effective than the mAb within grace period strategy which reflected real-world implementation. Both strategies showed large absolute effects against all-cause bronchiolitis, highlighting burden that test-negative designs may miss. Maternal RSVpreF’s key advantage is protection from birth, making it especially valuable where delayed mAbs are likely. As one of the first U.S. studies to directly compare these strategies, our findings contribute to individual, clinical, and programmatic decisions for infant RSV prevention.

## Data Availability

The data used in this study were licensed from Merative (MarketScan Commercial Claims and Encounters Database) under a data use agreement with Emory University. Researchers interested in accessing these data may do so by obtaining a license directly from Merative (https://www.merative.com).

## Acknowledgements

COI: SK has received institutional research support to conduct clinical trials from the NIH, CDC, Pfizer, Moderna, Sanofi, and Bavarian Nordic.

