## Supplemental Materials for "Comparative effectiveness of preventive strategies against medically-attended respiratory syncytial virus in U.S. infants during the first six months of life, 2023-2025"

Table S1. Specification of the target trial to compare the effectiveness of RSV preventive strategies against RSV hospitalization and medically-attended RSV

| **Protocol component** | **Target trial** | **Implementation with observational data** |
| --- | --- | --- |
| **Eligibility** | - Inclusion: Infants born during the 2023-2024 (October 1, 2023 through March 31, 2024) and 2024-2025 (October 1, 2024 through March 31, 2025) RSV seasons - Exclusion: Mother’s medical history during pregnancy is unknown or infant received palivizumab | - Inclusion: Same as target trial - Exclusion: Unable to be paired with mother’s claims record, not enrolled in insurance within 75 days from birth, or infant received palivizumab |
| **Intervention Strategies** | - Maternal receipt of RSVpreF ≥14 days prior to delivery or <14 days prior to delivery with mAb receipt within six months from birth - Infant receipt of mAbs within first week of life - Infant receipt of mAbs within six months from birth - No intervention as control | - Intervention groups are the same as target trial - Receipt of neither intervention for the control |
| **Assignment procedures** | - Randomized, unblinded | - Observed, use of inverse probability of treatment weights to mimic randomization |
| **Follow-up** | - From birth until the first of an outcome, loss to follow up, or end of follow-up at 6 months of age | - Same as target trial - Loss to follow up indicated by loss of insurance coverage |
| **Outcomes** | - Laboratory confirmed RSV hospitalization - Laboratory confirmed medically-attended RSV | - Hospitalization with RSV ICD-10 code - Outpatient or inpatient encounter with RSV ICD-10 code |
| **Causal Contrast** | - Cumulative incidence and hazard of the outcome from birth to six months by:  1) maternal RSVpreF vaccination versus control 2) mAb as intended versus control 3) mAb within grace period versus control 4) mAb within grace period versus mAb as intended 5) mAb as intended versus maternal RSVpreF 6) mAb within grace period versus maternal RSVpreF | - Same as target trial |

Abbreviations: mAb: long-acting monoclonal antibody; RSVpreF: Bivalent prefusion F vaccine

Table S2. Person time contributions for types of infants to each prevention strategy

| Type of infant | Contribution to the estimation of RSVpreF prevention strategy | Contribution to the estimation of mAb effectiveness | Contribution to the estimation under a control strategy |
| --- | --- | --- | --- |
| RSVpreF, delivery > 14 days after receipt and no mAb received after birth | All weeks of follow up from birth until RSV-associated outcome, six months of follow-up, or right-censoring event, whichever occurs first | None | None |
| RSVpreF, delivery > 14 days after receipt and mAb received after birth | All weeks of follow up from birth until RSV-associated outcome, six months of follow-up, right-censoring event, or mAb administration, whichever occurs first | None | None |
| RSVpreF, delivery ≤ 14 days after receipt and mAb received after birth | All weeks of follow up from birth until RSV-associated outcome, six months of follow-up, or right-censoring event, whichever occurs first | None | None |
| RSVpreF, delivery ≤ 14 days after receipt and no mAb received after birth | All weeks of follow up from birth until RSV-associated outcome, six months of follow-up, or right-censoring event, whichever occurs first | None | None |
| No RSVpreF and mAb received as intended (within first week of life) or within the grace period (within first six months of life) | None | All weeks of follow up from birth until RSV-associated outcome, six months of follow-up, or right-censoring event, whichever occurs first. | All weeks of follow up time from birth until RSV-associated outcome, mAb receipt, six months of follow-up, or right-censoring event, whichever occurs first. |
| No RSVpreF and mAb received outside of the as intended window (≥1 week from birth) | None | All weeks of follow up from birth until RSV-associated outcome, right-censoring event, or the end of the first week of life, whichever occurs first. | All weeks of follow up time from birth until RSV-associated outcome, mAb receipt, six months of follow-up, or right-censoring event, whichever occurs first. |
| No RSVpreF and mAb received outside of grace period (≥6 months from birth) | None | All weeks of follow up from birth until RSV-associated outcome, six months of follow-up, right-censoring event, whichever occurs first. | All weeks of follow up time from birth until RSV-associated outcome, mAb receipt, six months of follow-up, or right-censoring event, whichever occurs first. |
| No RSVpreF and mAb never received | None | All weeks of follow up time from birth until RSV-associated outcome, six months of follow-up, right censoring event, or the end of the first week of life for the as intended strategy, whichever occurs first. | All weeks of follow up from birth until RSV-associated outcome, six months of follow-up, or right-censoring event, whichever occurs first. |

Abbreviations: mAb: long-acting monoclonal antibody

Table S3. Descriptive characteristics by maternal RSVpreF timing relative to delivery and supplemental monoclonal antibody receipt

|  | Maternal RSVpreF ≥14 days from delivery | Maternal RSVpreF <14 days + mAb | Maternal RSVpreF <14 days + no mAb |
| --- | --- | --- | --- |
|  | N = 18,821^a^ | N = 229^a^ | N = 653^a^ |
| RSV Hospitalization | 35 (0.2%) | 0 (0%) | 6 (0.9%) |
| Medically-Attended RSV | 208 (1.1%) | 5 (2.2%) | 16 (2.5%) |
| Birth Season |  |  |  |
| 2023-2024 | 4804 (26%) | 56 (24%) | 202 (31%) |
| 2024-2025 | 14,017 (74%) | 173 (76%) | 451 (69%) |
| Birth Month |  |  |  |
| October | 2152 (11%) | 56 (24%) | 118 (18%) |
| November | 2859 (15%) | 47 (21%) | 139 (21%) |
| December | 3454 (18%) | 51 (22%) | 124 (19%) |
| January | 4219 (22%) | 51 (22%) | 159 (24%) |
| February | 3844 (20%) | 16 (7.0%) | 83 (13%) |
| March | 2293 (12%) | 3 (1.3%) | 30 (4.6) |
| Maternal Age | 32.79 (4.32) | 32.89 (4.62) | 33.25 (4.48) |
| Number of Prenatal Encounters | 4.51 (3.25) | 3.72 (2.87) | 3.81 (3.01) |
| Birth Region |  |  |  |
| Midwest | 5632 (30%) | 86 (38%) | 160 (25%) |
| Northeast | 3590 (19%) | 25 (11%) | 125 (19%) |
| South | 4946 (27%) | 65 (29%) | 196 (30%) |
| West | 4302 (23%) | 51 (22%) | 162 (25%) |
| Unknown | 351 | 2 | 10 |
| Preterm Birth | 1198 (6.4%) | 110 (48%) | 309 (47%) |
| Any Newborn Complication | 4580 (24%) | 145 (63%) | 379 (58%) |
| Maternal Comorbidity Affecting Pregnancy | 12,428 (66%) | 166 (72%) | 492 (75%) |
| Maternal Obstetric Complication | 9098 (48%) | 159 (69%) | 457 (70%) |

Abbreviations: mAb: long-acting monoclonal antibody; RSVpreF: Bivalent prefusion F vaccine

^a^n (%); Mean (Standard Deviation).

Table S4. Descriptive characteristics by monoclonal antibody timing

|  | mAb as intended (<1 week) | mAb within grace period but not as intended (≥1 week & < 6 months) |
| --- | --- | --- |
|  | N = 7227^e^ | N = 11,161^e^ |
| RSV Hospitalization | 11 (0.2%) | 43 (0.4%) |
| Medically-Attended RSV | 97 (1.3%) | 261 (2.3%) |
| Birth Season |  |  |
| 2023-2024 | 2838 (39%) | 6069 (54%) |
| 2024-2025 | 4389 (61%) | 5092 (46%) |
| Birth Month |  |  |
| October | 1221 (17%) | 4506 (40%) |
| November | 1382 (19%) | 2433 (22%) |
| December | 1207 (17%) | 1524 (14%) |
| January | 1234 (17%) | 1409 (13%) |
| February | 1100 (15%) | 814 (7.3%) |
| March | 1083 (15%) | 475 (4.3%) |
| Maternal Age | 32.25 (4.42) | 32.22 (4.50) |
| Number of Prenatal Encounters | 4.16 (3.17) | 4.12 (3.18) |
| Birth Region |  |  |
| Midwest | 1770 (25%) | 2494 (23%) |
| Northeast | 1140 (16%) | 1646 (15%) |
| South | 2827 (40%) | 4764 (43%) |
| West | 1336 (19%) | 2114 (19%) |
| Unknown | 154 | 143 |
| Preterm Birth | 653 (9.0%) | 1521 (14%) |
| Any Newborn Complication | 1806 (25%) | 3654 (33%) |
| Maternal Comorbidity Affecting Pregnancy | 4622 (64%) | 7121 (64%) |
| Maternal Obstetric Complication | 3370 (47%) | 5712 (51%) |

Abbreviations: mAb: long-acting monoclonal antibody

^a^n (%); Mean (Standard Deviation).

Table S5: Summary effectiveness (1-aHR)*100% with 95% confidence intervals of RSV-specific outcomes with diagnostic test ordered

|  | **Maternal RSVpreF vs control** | **mAb as intended vs control** | **mAb within grace period vs control** |
| --- | --- | --- | --- |
| **RSV hospitalization** | 84.3% (69.9% - 94.5%) | 79.0% (50.8% - 100.0%) | 77.8% (61.3% - 91.2%) |
| **Medically-attended RSV** | 66.6% (57.9% - 73.6%) | 61.8% (49.4% - 72.0%) | 54.9% (44.8% - 63.6%) |

Abbreviations: aHR: adjusted hazard ratio; mAb: long-acting monoclonal antibody; RSVpreF: Bivalent prefusion F vaccine

Table S6: Summary effectiveness ([1-aHR]*100%) of intervention strategy compared to control for bronchiolitis and all-cause acute respiratory illness

|  | Maternal RSVpreF vs control | mAb as intended vs control | mAb within grace period vs control |
| --- | --- | --- | --- |
| Bronchiolitis hospitalization | 32.3% (3.3% - 58.5%) | 13.0% (-36.4% - 57.6%) | 17.0% (-19.8% - 49.1%) |
| Season |  |  |  |
| 2023-2024 | 30.5% (-18.8% - 69.0%) | 4.9% (-74.0% – 66.0%) | 5.2% (-47.0% - 47.3%) |
| 2024-2025 | 34.4% (-3.6% - 60.6%) | 21.2 (-42.5% - 68.3%) | 29.9% (-24.7% - 69.3%) |
| Preterm |  |  |  |
| No | 29.7% (4.2% - 57.6%) | 6.2% (-55.6% - 57.5%) | 7.2% (-36.5% - 45.3%) |
| Yes | 33.9% (-57.9% - 90.6%) | 24.6% (-63.8% - 90.7%) | 46.2% (-12.0% - 87.7%) |
| Birth relative to season |  |  |  |
| Early (Oct – Dec) | 19.5% (-30.3% - 57.2%) | 53.9% (11.8% - 89.8%) | 45.2% (13.8% - 69.7%) |
| Late (Jan – Mar) | 39.6% (-4.3% - 72.7%) | -46.6 (-154.6% - 39.1%) | -25.1 (-111.4% - 43.7%) |
| Region |  |  |  |
| Midwest | 43.3% (-12.0% - 79.9%) | 0.7 (-102.7% - 73.3%) | 26.2% (39.8% - 76.9%) |
| Northeast | 58.4% (13.8% - 87.3%) | 23.6% (-110.4% - 92.9%) | 16.8% (-78.7% - 78.1%) |
| South | 9.8% (-65.5% - 64.9%) | 43.7% (-21.4% - 89.3%) | 17.7% (-48.4% - 63.7%) |
| West | 36.7% (-29.4% - 79.1%) | -24.9% (-176.3% - 100%) | 4.9% (-96.1% - 80.1%) |
| Medically-attended Bronchiolitis | 30.2% (20.4% - 38.8%) | 21.5% (8.7% - 33.2%) | 17.3% (6.0% - 26.9%) |
| Season |  |  |  |
| 2023-2024 | 37.0% (22.5% - 49.5%) | 32.7% (15.0% - 48.7%) | 18.4% (4.4% - 31.3%) |
| 2024-2025 | 21.9% (9.8% - 32.0%) | 10.3% (-9.0% - 27.7%) | 16.7% (1.2% - 30.5%) |
| Preterm |  |  |  |
| No | 29.2% (18.9% - 38.3%) | 23.3% (10.0% - 35.4%) | 18.7% (7.9% - 28.8%) |
| Yes | 35.3% (5.5% - 60.6%) | 0.2% (-51.6% - 40.4%) | 5.8% (-25.1% - 31.6%) |
| Birth relative to season |  |  |  |
| Early (Oct – Dec) | 31.5% (20.5% - 41.3%) | 29.3% (14.1% - 42.5%) | 25.2% (14.1% - 35.0%) |
| Late (Jan – Mar) | 16.4% (-1.5% - 31.5%) | 4.1% (-21.5% - 26.4%) | 0.3% (-23.3% - 20.2%) |
| Region |  |  |  |
| Midwest | 38.1% (22.7% - 50.7%) | 26.9% (0.8% - 48.8%) | 22.0% (2.2% - 40.0%) |
| Northeast | 27.6% (7.1% - 46.4%) | 15.4% (-19.2% - 45.5%) | 10.0% (-17.2% - 34.0%) |
| South | 22.8% (4.3% - 39.1%) | 22.0% (3.9% - 38.7%) | 13.6% (-2.6% - 26.7%) |
| West | 37.3% (18.7% - 52.3%) | 19.7% (-13.8% - 51.4%) | 27.6% (1.3% - 51.0%) |
| ARI hospitalization | 30.5% (6.6% - 50.1%) | 25.2% (-6.7% - 52.3%) | 23.3% (-1.0% - 45.8%) |
| Season |  |  |  |
| 2023-2024 | 32.7% (-8.0% - 61.9%) | 22.3 (-28.4% - 62.9%) | 16.5% (-18.8 - 46.0%) |
| 2024-2025 | 28.4% (-0.7% - 49.3%) | 28.9% (-11.2% - 62.1%) | 31.0% (-3.8% - 60.9%) |
| Preterm |  |  |  |
| No | 24.3% (-3.1% - 47.8%) | 20.0% (-18.5% - 55.2%) | 11.1% (-20.5% - 39.9%) |
| Yes | 42.6% (-8.9% - 79.6%) | 29.7% (-35.2% - 77.0%) | 57.3% (20.9% - 84.4%) |
| Birth relative to season |  |  |  |
| Early (Oct – Dec) | 21.2% (-13.5% - 49.1%) | 53.5% (21.4% - 79.5%) | 43.4% (17.5% - 62.6%) |
| Late (Jan – Mar) | 35.7% (1.4% - 61.9%) | -11.8% (-74.4% - 41.0%) | -3.5% (-53.9% - 41.1%) |
| Region |  |  |  |
| Midwest | 39.6% (-0.7% - 70.7%) | 13.1% (-55.2% - 68.6%) | 31.3% (-18.7% - 70.6%) |
| Northeast | 49.0% (10.3% - 78.1%) | 35.7% (-55.3% - 92.4%) | 24.7% (-45.2% - 75.2%) |
| South | 25.2% (-23.7% - 62.4%) | 37.9% (-16.1% - 75.8%) | 14.6% (-35.2% - 50.1%) |
| West | 17.5% (40.0% - 55.5%) | 11.3% (-78.9% - 83.9%) | 27.7% (-35.1% - 77.2%) |
| Medically-attended ARI | 12.6% (7.1% - 17.4%) | -2.5% (-9.3% - 3.9%) | -0.1% (-5.0% - 5.0%) |
| Season |  |  |  |
| 2023-2024 | 18.4% (10.3% - 25.7%) | -1.4% (-11.2% - 8.2%) | 0.2% (-6.9% - 7.0%) |
| 2024-2025 | 4.8% (-1.5% - 10.4%) | -3.7% (-13.1% - 4.9%) | -0.5% (-7.9% - 6.8%) |
| Preterm |  |  |  |
| No | 11.0% (5.7% - 16.1%) | 0.8% (06.2% - 7.1%) | 1.4% (-4.1% - 6.7%) |
| Yes | 27.6% (7.2% - 43.4%) | -41.4% (-70.2% - -12.8%) | -13.4% (-29.9% - 1.1%) |
| Birth relative to season |  |  |  |
| Early (Oct – Dec) | 14.0% (6.8% - 20.0%) | 5.0% (-2.9% - 12.6%) | 7.1% (1.9% - 12.5%) |
| Late (Jan – Mar) | 4.2% (-4.5% - 12.0%) | -14.4% (-25.7% - -3.2%) | -11.6% (-20.9% - -2.5%) |
| Region |  |  |  |
| Midwest | 10.8% (1.5% - 18.9%) | 2.5% (-10.5% - 14.4%) | 3.9% (-6.5% - 13.4%) |
| Northeast | 22.8% (12.9% - 31.7%) | -2.1% (-19.7% - 12.8%) | 1.5% (-11.6% - 13.6%) |
| South | 9.4% (0.5% - 17.6%) | -3.6% (-13.8% - 5.8%) | -3.2% (-11.2% - 3.9%) |
| West | 10.5% (-5.9% - 22.3%) | -7.8% (-26.1% - 8.7%) | -0.6% (-13.6% - 11.6%) |

Abbreviations: aHR: adjusted hazard ratio; mAb: long-acting monoclonal antibody; RSVpreF: Bivalent prefusion F vaccine; ARI: acute respiratory illness

Table S7: Summary adjusted hazard ratio with 95% confidence intervals among first five weeks of follow-up versus after five weeks

|  | Maternal RSVpreF vs control | mAb as intended vs control | mAb within grace period vs control | mAb within grace period vs mAb as intended | mAb as intended vs maternal RSVpreF | mAb within grace period vs maternal RSVpreF |
| --- | --- | --- | --- | --- | --- | --- |
| RSV hospitalization | | | | | | |
| ≤5 weeks | 0.2 (0.1 – 0.3) | 0.2 (0.1 – 0.4) | 0.4 (0.2 – 0.5) | 1.8 (1.1 – 5.7) | 1.10 (0.2 – 3.4) | 2.0 (1.0 – 4.9) |
| >5 weeks | 0.2 (0.1 – 0.2) | 0.1 (0.0 – 0.3) | 0.1 (0.0 – 0.2) | 0.8 (0.5 – 4.1) | 0.8 (0.1 – 2.0) | 0.7 (0.2 – 1.5) |
| Medically-attended RSV | | | | | | |
| ≤5 weeks | 0.4 (0.2 – 0.5) | 0.4 (0.3 – 0.6) | 0.6 (0.4 – 0.7) | 1.3 (1.0 – 1.7) | 1.2 (0.7 – 2.3) | 1.6 (1.0 – 2.7) |
| >5 weeks | 0.3 (0.3 – 0.4) | 0.3 (0.2 – 0.4) | 0.4 (0.3 – 0.5) | 1.2 (1.0 – 1.5) | 0.9 (0.6 – 1.2) | 1.1 (0.8 – 1.4) |

Abbreviations: mAb: long-acting monoclonal antibody; RSVpreF: Bivalent prefusion F vaccine; ARI: acute respiratory illness

Table S8: Distribution of maternal characteristics among RSVpreF recipients by season

| Characteristic | 2023-2024  N=5,062^a^ | 2024-2025  N=14,641^a^ |
| --- | --- | --- |
| RSV Hospitalization | 22 (0.4%) | 40 (0.3%) |
| Medically-Attended RSV | 221 (4.4%) | 216 (1.5%) |
| Month RSVPreF Received |  |  |
| August | 0 (0%) | 63 (0.4%) |
| September | 22 (0.4%) | 2,510 (17%) |
| October | 319 (6.3%) | 3,178 (22%) |
| November | 1,025 (20%) | 2,554 (17%) |
| December | 1,426 (28%) | 2,910 (20%) |
| January | 1,889 (37%) | 3,017 (21%) |
| February | 354 (7.0%) | 382 (2.6%) |
| March | 27 (0.5%) | 27 (0.2%) |
| Maternal Age | 33.07 (4.23) | 32.72 (4.36) |
| Number of Prenatal Encounters | 4.38 (3.23) | 4.51 (3.24) |
| Birth Region |  |  |
| Midwest | 1,567 (32%) | 4,311 (30%) |
| Northeast | 969 (19%) | 2,771 (19%) |
| South | 1,027 (21%) | 4,180 (29%) |
| West | 1,411 (28%) | 3,104 (22%) |
| Unknown | 88 | 275 |
| Preterm Birth | 448 (8.9%) | 1,169 (8.0%) |
| Maternal Cardiovascular Conditions | 1,438 (28%) | 4,222 (29%) |
| Gestational Diabetes | 714 (14%) | 2,165 (15%) |
| Maternal Obesity | 1,240 (24%) | 3,970 (27%) |
| Maternal Liver Conditions | 120 (2.4%) | 305 (2.1%) |
| Maternal Hematological Conditions | 278 (5.5%) | 830 (5.7%) |
| Maternal Immunocompromising Conditions | 227 (4.5%) | 706 (4.8%) |

^a^n (%); Mean (Standard Deviation)

Abbreviations: RSVpreF: Bivalent prefusion F vaccine

Figure S1. Cumulative probability of mAb receipt over age in weeks by season

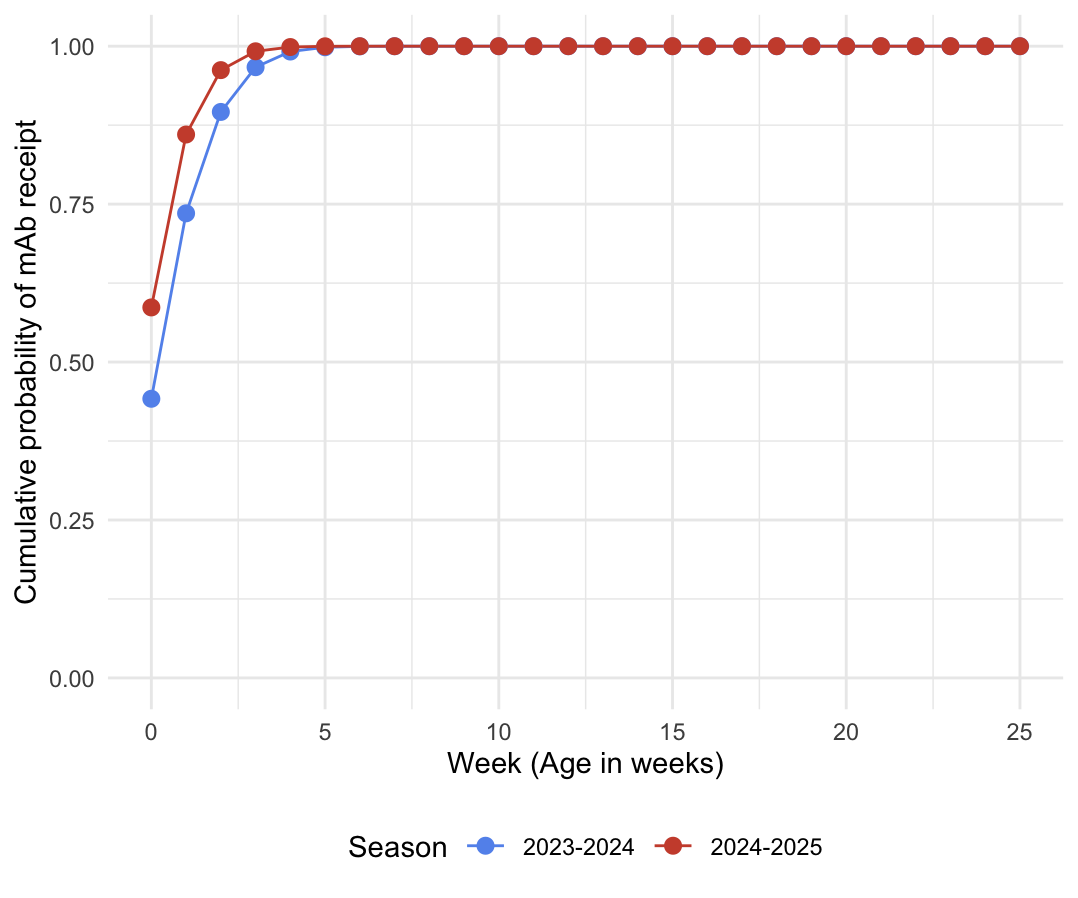

Abbreviations: mAb: long-acting monoclonal antibody

Figure S2. Risk difference with 95% confidence intervals of RSV hospitalization and medically-attended RSV over time by intervention strategy contrast

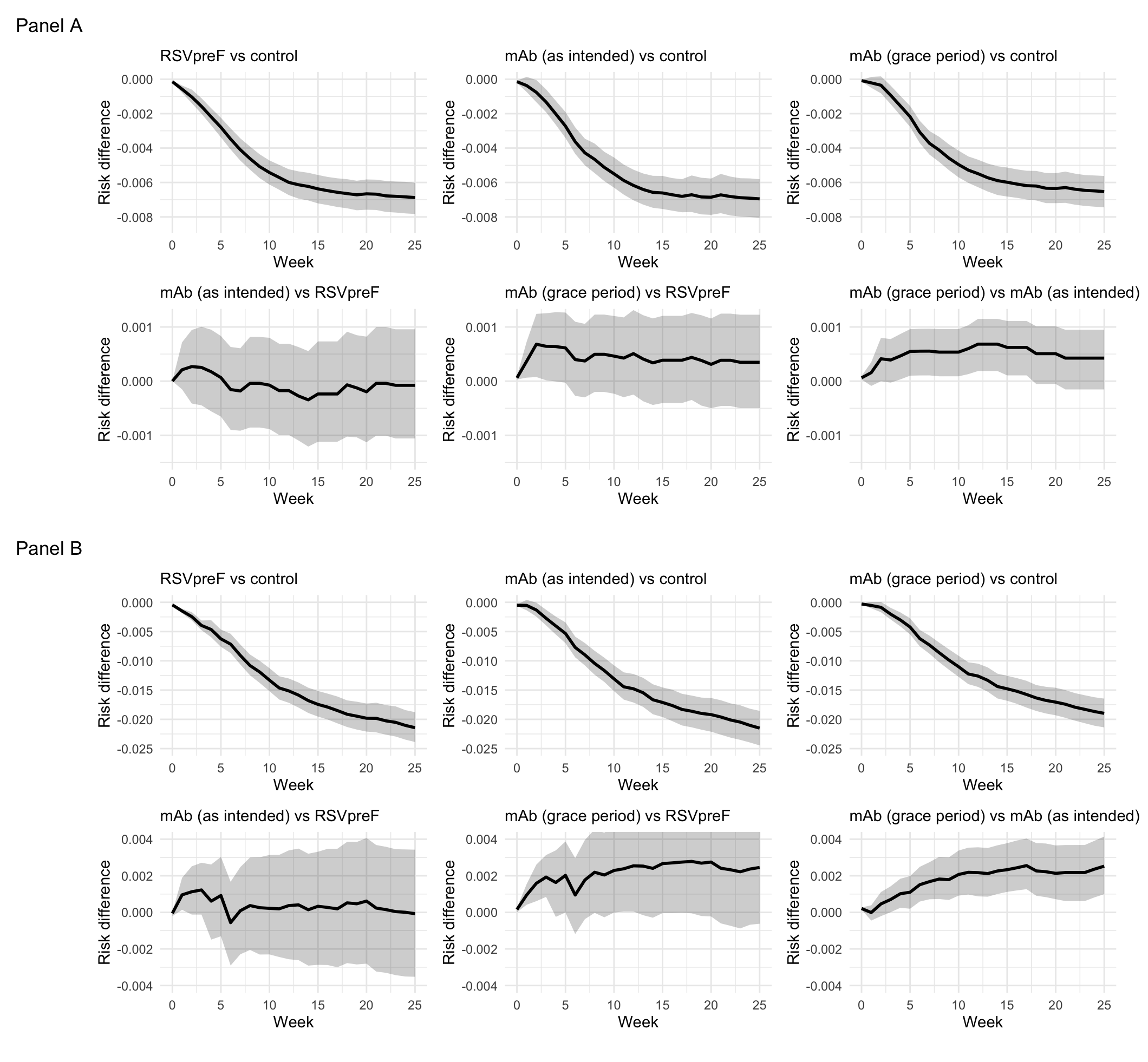

Caption: Panel A: Risk difference of RSV hospitalization; Panel B: Risk difference of medically-attended RSV

Abbreviations: mAb: long-acting monoclonal antibody; RSVpreF: Bivalent prefusion F vaccine

Figure S3. Strategy specific cumulative incidence for RSV hospitalization and medically-attended RSV by season

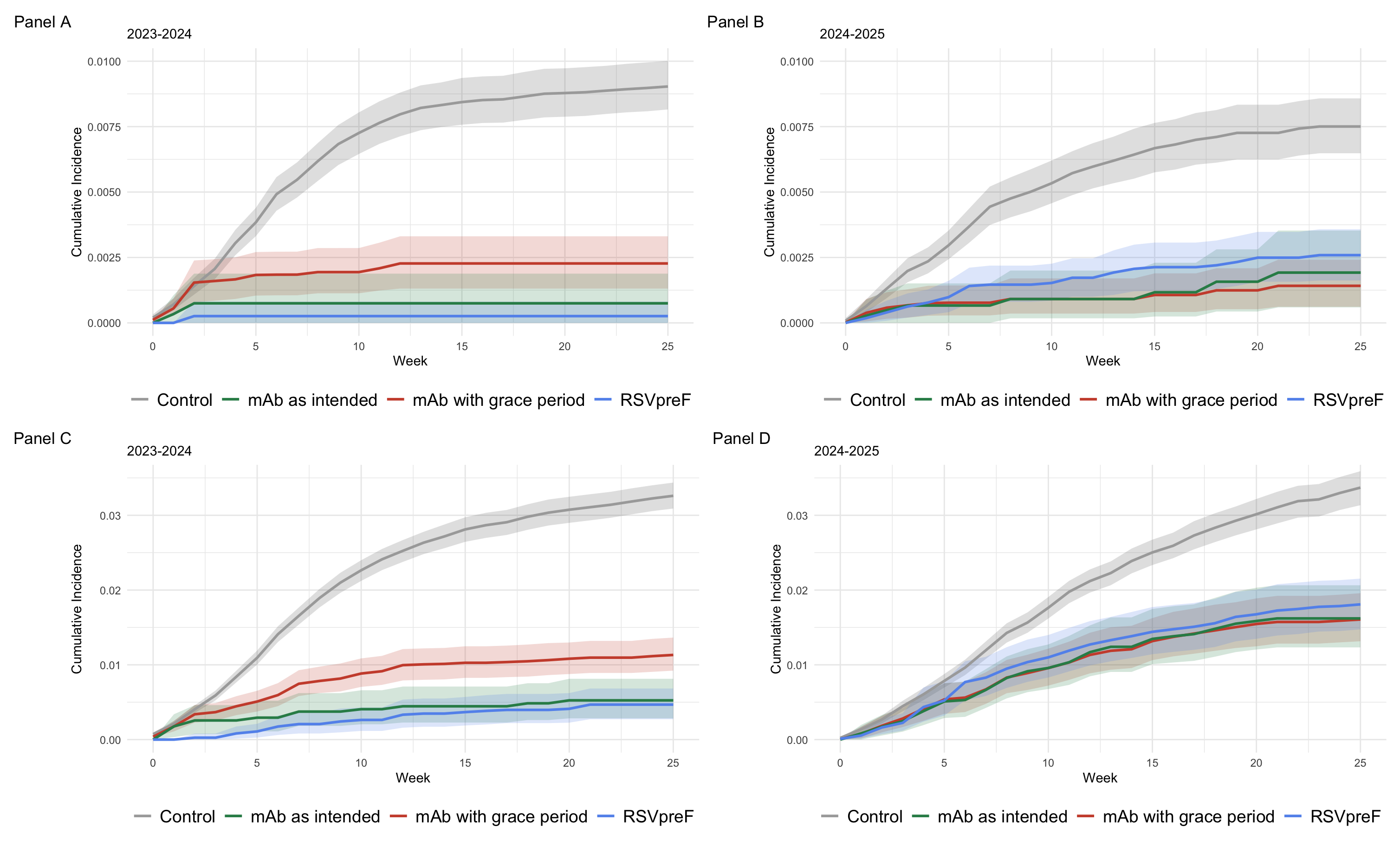

Caption: Panel A: RSV hospitalization 2023-2024; Panel B: RSV hospitalization 2024-2025; Panel C: medically-attended RSV 2023-2024; Panel D: medically-attended RSV 2024-2025

Abbreviations: mAb: long-acting monoclonal antibody; RSVpreF: Bivalent prefusion F vaccine

Appendix 1: Birth and delivery related ICD-10 codes to identify newborn infants and linked mothers in MarketScan

| Code | Type | Description |
| --- | --- | --- |
| Z38 (category) | ICD-10-CM | Liveborn infants according to place of birth and type of delivery |
| Z37 (category) | ICD-10-CM | Outcome of delivery (mother's record) — includes single live birth (Z37.0), twin live births (Z37.1/.2/.3), and other multiple birth outcomes |
| P08.21 | ICD-10-CM | Post-term newborn |
| P08.22 | ICD-10-CM | Prolonged gestation of newborn |
| P07.00 | ICD-10-CM | Extremely low birth weight newborn, unspecified weight |
| P07.01 | ICD-10-CM | Extremely low birth weight newborn, less than 500 grams |
| P07.02 | ICD-10-CM | Extremely low birth weight newborn, 500–749 grams |
| P07.03 | ICD-10-CM | Extremely low birth weight newborn, 750–999 grams |
| P07.10 | ICD-10-CM | Other low birth weight newborn, unspecified weight |
| P07.14 | ICD-10-CM | Other low birth weight newborn, 1000–1249 grams |
| P07.15 | ICD-10-CM | Other low birth weight newborn, 1250–1499 grams |
| P07.16 | ICD-10-CM | Other low birth weight newborn, 1500–1749 grams |
| P07.17 | ICD-10-CM | Other low birth weight newborn, 1750–1999 grams |
| P07.18 | ICD-10-CM | Other low birth weight newborn, 2000–2499 grams |
| P07.21 | ICD-10-CM | Extreme immaturity of newborn, gestational age less than 23 completed weeks |
| P07.22 | ICD-10-CM | Extreme immaturity of newborn, gestational age 23 completed weeks |
| P07.23 | ICD-10-CM | Extreme immaturity of newborn, gestational age 24 completed weeks |
| P07.24 | ICD-10-CM | Extreme immaturity of newborn, gestational age 25 completed weeks |
| P07.25 | ICD-10-CM | Extreme immaturity of newborn, gestational age 26 completed weeks |
| P07.26 | ICD-10-CM | Extreme immaturity of newborn, gestational age 27 completed weeks |
| P07.30 | ICD-10-CM | Preterm newborn, unspecified weeks of gestation |
| P07.31 | ICD-10-CM | Preterm newborn, gestational age 28 completed weeks |
| P07.32 | ICD-10-CM | Preterm newborn, gestational age 29 completed weeks |
| P07.33 | ICD-10-CM | Preterm newborn, gestational age 30 completed weeks |
| P07.34 | ICD-10-CM | Preterm newborn, gestational age 31 completed weeks |
| P07.35 | ICD-10-CM | Preterm newborn, gestational age 32 completed weeks |
| P07.36 | ICD-10-CM | Preterm newborn, gestational age 33 completed weeks |
| P07.37 | ICD-10-CM | Preterm newborn, gestational age 34 completed weeks |
| P07.38 | ICD-10-CM | Preterm newborn, gestational age 35 completed weeks |
| P07.39 | ICD-10-CM | Preterm newborn, gestational age 36 completed weeks |
| P01.5 | ICD-10-CM | Newborn affected by multiple pregnancy |
| O80 | ICD-10-CM | Encounter for full-term uncomplicated delivery |
| O82 | ICD-10-CM | Encounter for cesarean delivery without indication |
| 59409 | CPT | Vaginal delivery only (with or without episiotomy and/or forceps) |
| 59410 | CPT | Vaginal delivery only, including postpartum care |
| 59514 | CPT | Cesarean delivery only |
| 59515 | CPT | Cesarean delivery only, including postpartum care |
| 59612 | CPT | Vaginal delivery only, after previous cesarean delivery |
| 59614 | CPT | Vaginal delivery only, after previous cesarean delivery, including postpartum care |
| 59620 | CPT | Cesarean delivery only, following attempted vaginal delivery after previous cesarean delivery |
| 59622 | CPT | Cesarean delivery only, following attempted vaginal delivery after previous cesarean delivery, including postpartum care |
| 765 | MS-DRG | Cesarean section with CC/MCC |
| 766 | MS-DRG | Cesarean section without CC/MCC |
| 767 | MS-DRG | Vaginal delivery with sterilization and/or D&C |
| 768 | MS-DRG | Vaginal delivery with O.R. procedure except sterilization and/or D&C |
| 774 | MS-DRG | Vaginal delivery with complicating diagnoses |
| 775 | MS-DRG | Vaginal delivery without complicating diagnoses |
| 790 | MS-DRG | Extreme immaturity or respiratory distress syndrome, neonate |
| 791 | MS-DRG | Prematurity with major problems |
| 792 | MS-DRG | Prematurity without major problems |
| 793 | MS-DRG | Full-term neonate with major problems |
| 795 | MS-DRG | Normal newborn |

Appendix 2: Current procedural terminology codes for identifying RSV preventive products

| Code | Type | Description |
| --- | --- | --- |
| 90678 | CPT | Respiratory syncytial virus vaccine, preF, subunit, bivalent, for intramuscular use (maternal RSVpreF vaccine, e.g., Abrysvo, given during pregnancy) |
| 90380 | CPT | Respiratory syncytial virus, monoclonal antibody, seasonal dose; 0.5 mL dosage, for intramuscular use (nirsevimab, e.g., Beyfortus) |
| 90381 | CPT | Respiratory syncytial virus, monoclonal antibody, seasonal dose; 1 mL dosage, for intramuscular use (nirsevimab, e.g., Beyfortus) |
| 96380 | CPT | Administration of respiratory syncytial virus, monoclonal antibody, seasonal dose by intramuscular injection, with counseling by physician or other qualified health care professional |
| 96381 | CPT | Administration of respiratory syncytial virus, monoclonal antibody, seasonal dose by intramuscular injection, without counseling by physician or other qualified health care professional |

Appendix 3: Acute respiratory ICD-10 codes

| Code | Description |
| --- | --- |
| A37.00 | Whooping cough due to Bordetella pertussis, without pneumonia |
| A37.01 | Whooping cough due to Bordetella pertussis, with pneumonia |
| A37.10 | Whooping cough due to Bordetella parapertussis, without pneumonia |
| A37.11 | Whooping cough due to Bordetella parapertussis, with pneumonia |
| A37.80 | Whooping cough due to other Bordetella species, without pneumonia |
| A37.81 | Whooping cough due to other Bordetella species, with pneumonia |
| A37.90 | Whooping cough, unspecified species, without pneumonia |
| A37.91 | Whooping cough, unspecified species, with pneumonia |
| B25.0 | Cytomegaloviral pneumonitis |
| B97.4 | Respiratory syncytial virus as the cause of diseases classified elsewhere |
| J00 | Acute nasopharyngitis [common cold] |
| J01.00 | Acute maxillary sinusitis, unspecified |
| J01.01 | Acute recurrent maxillary sinusitis |
| J01.10 | Acute frontal sinusitis, unspecified |
| J01.11 | Acute recurrent frontal sinusitis |
| J01.20 | Acute ethmoidal sinusitis, unspecified |
| J01.21 | Acute recurrent ethmoidal sinusitis |
| J01.30 | Acute sphenoidal sinusitis, unspecified |
| J01.31 | Acute recurrent sphenoidal sinusitis |
| J01.40 | Acute pansinusitis, unspecified |
| J01.41 | Acute recurrent pansinusitis |
| J01.80 | Other acute sinusitis |
| J01.81 | Other acute recurrent sinusitis |
| J01.90 | Acute sinusitis, unspecified |
| J01.91 | Acute recurrent sinusitis, unspecified |
| J02.0 | Streptococcal pharyngitis |
| J02.8 | Acute pharyngitis due to other specified organisms |
| J02.9 | Acute pharyngitis, unspecified |
| J03.00 | Acute streptococcal tonsillitis, unspecified |
| J03.01 | Acute recurrent streptococcal tonsillitis |
| J03.80 | Acute tonsillitis due to other specified organisms |
| J03.81 | Acute recurrent tonsillitis due to other specified organisms |
| J03.90 | Acute tonsillitis, unspecified |
| J03.91 | Acute recurrent tonsillitis, unspecified |
| J04.0 | Acute laryngitis |
| J04.10 | Acute tracheitis without obstruction |
| J04.11 | Acute tracheitis with obstruction |
| J04.2 | Acute laryngotracheitis |
| J04.30 | Supraglottitis, unspecified, without obstruction |
| J04.31 | Supraglottitis, unspecified, with obstruction |
| J05.0 | Acute obstructive laryngitis [croup] |
| J05.10 | Acute epiglottitis without obstruction |
| J05.11 | Acute epiglottitis with obstruction |
| J06.0 | Acute laryngopharyngitis |
| J06.9 | Acute upper respiratory infection, unspecified |
| J09.X1 | Influenza due to identified novel influenza A virus with pneumonia |
| J09.X2 | Influenza due to identified novel influenza A virus with other respiratory manifestations |
| J09.X3 | Influenza due to identified novel influenza A virus with gastrointestinal manifestations |
| J09.X9 | Influenza due to identified novel influenza A virus with other manifestations |
| J10.00 | Influenza due to other identified influenza virus with unspecified type of pneumonia |
| J10.01 | Influenza due to other identified influenza virus with the same other identified influenza virus pneumonia |
| J10.08 | Influenza due to other identified influenza virus with other specified pneumonia |
| J10.1 | Influenza due to other identified influenza virus with other respiratory manifestations |
| J10.2 | Influenza due to other identified influenza virus with gastrointestinal manifestations |
| J10.81 | Influenza due to other identified influenza virus with encephalopathy |
| J10.82 | Influenza due to other identified influenza virus with myocarditis |
| J10.83 | Influenza due to other identified influenza virus with otitis media |
| J10.89 | Influenza due to other identified influenza virus with other manifestations |
| J11.00 | Influenza due to unidentified influenza virus with unspecified type of pneumonia |
| J11.08 | Influenza due to unidentified influenza virus with specified pneumonia |
| J11.1 | Influenza due to unidentified influenza virus with other respiratory manifestations |
| J11.2 | Influenza due to unidentified influenza virus with gastrointestinal manifestations |
| J11.81 | Influenza due to unidentified influenza virus with encephalopathy |
| J11.82 | Influenza due to unidentified influenza virus with myocarditis |
| J11.83 | Influenza due to unidentified influenza virus with otitis media |
| J11.89 | Influenza due to unidentified influenza virus with other manifestations |
| J12.0 | Adenoviral pneumonia |
| J12.1 | Respiratory syncytial virus pneumonia |
| J12.2 | Parainfluenza virus pneumonia |
| J12.3 | Human metapneumovirus pneumonia |
| J12.81 | Pneumonia due to SARS-associated coronavirus |
| J12.89 | Other viral pneumonia |
| J12.9 | Viral pneumonia, unspecified |
| J13 | Pneumonia due to Streptococcus pneumoniae |
| J14 | Pneumonia due to Hemophilus influenzae |
| J15.0 | Pneumonia due to Klebsiella pneumoniae |
| J15.1 | Pneumonia due to Pseudomonas |
| J15.20 | Pneumonia due to staphylococcus, unspecified |
| J15.211 | Pneumonia due to Methicillin susceptible Staphylococcus aureus |
| J15.212 | Pneumonia due to Methicillin resistant Staphylococcus aureus |
| J15.29 | Pneumonia due to other staphylococcus |
| J15.3 | Pneumonia due to streptococcus, group B |
| J15.4 | Pneumonia due to other streptococci |
| J15.5 | Pneumonia due to Escherichia coli |
| J15.6 | Pneumonia due to other Gram-negative bacteria (non-billable parent code) |
| J15.61 | Pneumonia due to Acinetobacter baumannii |
| J15.69 | Pneumonia due to other Gram-negative bacteria |
| J15.7 | Pneumonia due to Mycoplasma pneumoniae |
| J15.8 | Pneumonia due to other specified bacteria |
| J15.9 | Unspecified bacterial pneumonia |
| J16.0 | Chlamydial pneumonia |
| J16.8 | Pneumonia due to other specified infectious organisms |
| J17 | Pneumonia in diseases classified elsewhere |
| J18.0 | Bronchopneumonia, unspecified organism |
| J18.1 | Lobar pneumonia, unspecified organism |
| J18.2 | Hypostatic pneumonia, unspecified organism |
| J18.8 | Other pneumonia, unspecified organism |
| J18.9 | Pneumonia, unspecified organism |
| J20.0 | Acute bronchitis due to Mycoplasma pneumoniae |
| J20.1 | Acute bronchitis due to Hemophilus influenzae |
| J20.2 | Acute bronchitis due to streptococcus |
| J20.3 | Acute bronchitis due to coxsackievirus |
| J20.4 | Acute bronchitis due to parainfluenza virus |
| J20.5 | Acute bronchitis due to respiratory syncytial virus |
| J20.6 | Acute bronchitis due to rhinovirus |
| J20.7 | Acute bronchitis due to echovirus |
| J20.8 | Acute bronchitis due to other specified organisms |
| J20.9 | Acute bronchitis, unspecified |
| J21.0 | Acute bronchiolitis due to respiratory syncytial virus |
| J21.1 | Acute bronchiolitis due to human metapneumovirus |
| J21.8 | Acute bronchiolitis due to other specified organisms |
| J21.9 | Acute bronchiolitis, unspecified |
| J22 | Unspecified acute lower respiratory infection |
| J39.8 | Other specified diseases of upper respiratory tract |
| J39.9 | Disease of upper respiratory tract, unspecified |
| J40 | Bronchitis, not specified as acute or chronic |
| J45.20 | Mild intermittent asthma, uncomplicated |
| J45.21 | Mild intermittent asthma with (acute) exacerbation |
| J45.22 | Mild intermittent asthma with status asthmaticus |
| J45.30 | Mild persistent asthma, uncomplicated |
| J45.31 | Mild persistent asthma with (acute) exacerbation |
| J45.32 | Mild persistent asthma with status asthmaticus |
| J45.40 | Moderate persistent asthma, uncomplicated |
| J45.41 | Moderate persistent asthma with (acute) exacerbation |
| J45.42 | Moderate persistent asthma with status asthmaticus |
| J45.50 | Severe persistent asthma, uncomplicated |
| J45.51 | Severe persistent asthma with (acute) exacerbation |
| J45.52 | Severe persistent asthma with status asthmaticus |
| J45.901 | Unspecified asthma with (acute) exacerbation |
| J45.902 | Unspecified asthma with status asthmaticus |
| J45.909 | Unspecified asthma, uncomplicated |
| J45.990 | Exercise induced bronchospasm |
| J45.991 | Cough variant asthma |
| J45.998 | Other asthma |
| J41.0 | Simple chronic bronchitis |
| J41.1 | Mucopurulent chronic bronchitis |
| J41.8 | Mixed simple and mucopurulent chronic bronchitis |
| J42 | Unspecified chronic bronchitis |
| J43.0 | Unilateral pulmonary emphysema [MacLeod's syndrome] |
| J43.1 | Panlobular emphysema |
| J43.2 | Centrilobular emphysema |
| J43.8 | Other emphysema |
| J43.9 | Emphysema, unspecified |
| J44.0 | Chronic obstructive pulmonary disease with (acute) lower respiratory infection |
| J47.0 | Bronchiectasis with acute lower respiratory infection |
| J47.1 | Bronchiectasis with (acute) exacerbation |

Appendix 4: CPT codes to identify RSV diagnostic test ordered

| Code | Type | Description |
| --- | --- | --- |
| 87280 | CPT | Infectious agent antigen detection by immunofluorescent technique; respiratory syncytial virus |
| 87420 | CPT | Infectious agent antigen detection by immunoassay technique (e.g., EIA, ELISA, FIA, IMCA), qualitative or semiquantitative; respiratory syncytial virus |
| 87807 | CPT | Infectious agent antigen detection by immunoassay with direct optical (visual) observation; respiratory syncytial virus |
| 86756 | CPT | Antibody; respiratory syncytial virus |
| 87634 | CPT | Infectious agent detection by nucleic acid (DNA or RNA); respiratory syncytial virus, amplified probe technique |
| 0202U | PLA (CPT) | Infectious disease (bacterial or viral respiratory tract infection), pathogen-specific nucleic acid (DNA or RNA), 22 targets including SARS-CoV-2, qualitative RT-PCR, nasopharyngeal swab (BioFire FilmArray Respiratory Panel 2.1) |
| 87631 | CPT | Infectious agent detection by nucleic acid (DNA or RNA); respiratory virus, multiplex reverse transcription and amplified probe technique, multiple types/subtypes, 3–5 targets |
| 87632 | CPT | Infectious agent detection by nucleic acid (DNA or RNA); respiratory virus, multiplex reverse transcription and amplified probe technique, multiple types/subtypes, 6–11 targets |
| 87633 | CPT | Infectious agent detection by nucleic acid (DNA or RNA); respiratory virus, multiplex reverse transcription and amplified probe technique, multiple types/subtypes, 12–25 targets |
| 87637 | CPT | Infectious agent detection by nucleic acid (DNA or RNA); SARS-CoV-2, influenza virus types A and B, and respiratory syncytial virus, multiplex amplified probe technique |
| 0241U | PLA (CPT) | Infectious disease (viral respiratory tract infection), pathogen-specific RNA, 4 targets (SARS-CoV-2, influenza A, influenza B, RSV), upper respiratory specimen, each pathogen reported as detected/not detected |

Appendix 5: ICD-10 codes for maternal comorbidities affecting pregnancy

| Code | Description |
| --- | --- |
| O10 (category) | Pre-existing hypertension complicating pregnancy, childbirth and the puerperium |
| O11 (category) | Pre-existing hypertension with pre-eclampsia |
| O12 (category) | Gestational edema and proteinuria without hypertension |
| O13 (category) | Gestational [pregnancy-induced] hypertension without significant proteinuria |
| O14 (category) | Pre-eclampsia |
| O15 (category) | Eclampsia |
| O16 (category) | Unspecified maternal hypertension |
| O24 (category) | Diabetes mellitus in pregnancy, childbirth, and the puerperium |
| O99.21 | Obesity complicating pregnancy, childbirth, and the puerperium (non-billable parent; matches billable subcodes O99.210–O99.215) |
| O99.5 (category) | Diseases of the respiratory system complicating pregnancy, childbirth and the puerperium |
| O26.6 (category) | Liver and biliary tract disorders in pregnancy, childbirth and the puerperium |
| O99.0 (category) | Anemia complicating pregnancy, childbirth and the puerperium |
| O99.1 (category) | Other diseases of the blood and blood-forming organs and certain disorders involving the immune mechanism complicating pregnancy, childbirth and the puerperium |
| O98.7 (category) | Human immunodeficiency virus [HIV] disease complicating pregnancy, childbirth and the puerperium |
| O9A (category) | Maternal malignant neoplasms, traumatic injuries and abuse classifiable elsewhere but complicating pregnancy, childbirth, and the puerperium |
| D80 (category) | Immunodeficiency with predominantly antibody defects |
| D81 (category) | Combined immunodeficiencies |
| D82 (category) | Immunodeficiency associated with other major defects |
| D83 (category) | Common variable immunodeficiency |
| D84 (category) | Other immunodeficiencies |
| D86 (category) | Sarcoidosis |
| D89 (category) | Other disorders involving the immune mechanism, not elsewhere classified |
| M35.9 | Systemic involvement of connective tissue, unspecified (systemic autoimmune/collagen-vascular disease NOS) |
| D71 (category) | Functional disorders of polymorphonuclear neutrophils |
| B20 (category) | Human immunodeficiency virus [HIV] disease |
| D70 (category) | Neutropenia |
| Z79.5x | Long term (current) use of steroids (Z79.51 inhaled, Z79.52 systemic) |
| Z94 (category) | Transplanted organ and tissue status |
| Q89.01 | Asplenia (congenital) |

Appendix 6: ICD-10 codes for newborn and obstetric complications

| Code | Description |
| --- | --- |
| P02.4 | Newborn affected by prolapsed cord |
| P02.5 | Newborn affected by other compression of umbilical cord |
| P02.6 | Newborn affected by other and unspecified conditions of umbilical cord |
| P51 (category) | Umbilical hemorrhage of newborn |
| Q20 (category) | Congenital malformations of cardiac chambers and connections |
| Q21 (category) | Congenital malformations of cardiac septa |
| Q22 (category) | Congenital malformations of pulmonary and tricuspid valves |
| Q23 (category) | Congenital malformations of aortic and mitral valves |
| Q24 (category) | Other congenital malformations of heart |
| Q25 (category) | Congenital malformations of great arteries |
| Q26 (category) | Congenital malformations of great veins |
| Q27 (category) | Other congenital malformations of peripheral vascular system |
| P29 (category) | Cardiovascular disorders originating in the perinatal period |
| P22 (category) | Respiratory distress of newborn |
| P25 (category) | Interstitial emphysema and related conditions originating in the perinatal period |
| P26 (category) | Pulmonary hemorrhage originating in the perinatal period |
| P27 (category) | Chronic respiratory disease originating in the perinatal period (e.g., bronchopulmonary dysplasia) |
| P28 (category) | Other respiratory conditions of newborn (e.g., atelectasis, neonatal apnea) |
| Q33 (category) | Congenital malformations of lung |
| P05 (category) | Disorders of newborn related to slow fetal growth and fetal malnutrition |
| P07 (category) | Disorders of newborn related to short gestation and low birth weight, not elsewhere classified |
| E84.0 | Cystic fibrosis with pulmonary manifestations |
| P94 (category) | Disorders of muscle tone of newborn |
| P20 (category) | Intrauterine hypoxia |
| P21 (category) | Birth asphyxia |
| O32 (category) | Maternal care for malpresentation of fetus (decision for cesarean delivery made before onset of labor) |
| O64 (category) | Obstructed labor due to malposition and malpresentation of fetus |
| O69 (category) | Labor and delivery complicated by umbilical cord complications (e.g., cord prolapse, cord around neck, cord entanglement) |
| O45 (category) | Premature separation of placenta [abruptio placentae] |
| O67 (category) | Labor and delivery complicated by intrapartum hemorrhage, not elsewhere classified |
| O14 (category) | Pre-eclampsia |
| O36.5 (category) | Maternal care for known or suspected poor fetal growth |
| O60 (category) | Preterm labor |
| D69 (category) | Purpura and other hemorrhagic conditions |
| D58 (category) | Other hereditary hemolytic anemias |
| D59 (category) | Acquired hemolytic anemia |
